# Age-related impacts of dementia status and subtype on accelerometry measures in older adults from the National Health and Aging Trends Study

**DOI:** 10.1101/2024.05.09.24307083

**Authors:** Karl Brown, Andrew Shutes-David, Sarah Payne, Adrienne Jankowski, Katie Wilson, Edmund Seto, Debby Tsuang

## Abstract

It is likely that age, dementia status, and dementia subtype all contribute to differences in activity levels among older adults and that accelerometer devices can help us better understand these relationships. To that end, this study analyzes accelerometry data from individuals with dementia (n=104) and individuals without dementia (n=634) who participated in the National Health and Aging Trends Study (NHATS), as well as from individuals with dementia with Lewy bodies (DLB; n=12) and Alzheimer’s disease (AD; n=8) who participated in a small pilot study. In the NHATS cohort, individuals with dementia had lower daily activity counts (t = -5.449, p < 0.001), a shorter active bout length (t = -5.283, p < 0.001), and a longer resting bout length (t = 2.210, p = 0.032) at the mean age of 79 than individuals without dementia at that same age. Differences in data collection and processing prevented direct comparisons between the cohorts, but parallel analyses revealed no statistically significant differences between AD and DLB across these three measures in the smaller cohort. Studies with larger samples of subtyped individuals with dementia will be necessary to detect clinically meaningful differences, but the patterns observed in the small cohort suggest that individuals with DLB are less active, exhibit a shorter active bout length, and exhibit a longer resting bout length than individuals with AD. Taken together, these findings highlight the potential of accelerometers to gather objective activity data that could be used to aid in the early identification of dementia and, potentially, the differentiation of dementia subtypes.

## Introduction

The importance of physical activity (PA) as a modifiable risk factor for cardiovascular disease, cancer, diabetes, and other causes of mortality (World Health Organization, 2020) has become much clearer over the last 30 years, in part due to the emergence of accelerometry-based activity monitors that can capture more accurate PA patterns than self-report measures. Over this time, we have also come to see a clearer relationship between PA and dementia. For example, traditional non-accelerometry studies have established that individuals with dementia are less active than age- and sex-matched controls (Barnes & Yaffe, 2011; Petermann-Rocha et al., 2021; Taylor et al., 2019; Zhu et al., 2017) and that individuals with dementia experience a variety of symptoms that affect their daily PA patterns, including sundowning (i.e., the worsening of neuropsychiatric symptoms in the late afternoon or early evening), sleep disturbances at night, and sleepiness during the day (Canevelli et al., 2016; Grace, Walker, & McKeith, 2000). By using accelerometry, other studies have shown that lower levels of daily activity, greater prolonged sedentary time, and less stable 24-hour activity patterns are associated with lower levels of cognition in older adults (Buchman, Wilson, & Bennett, 2008; Chen et al., 2022; Farina, Tabet, & Rusted, 2014; Luik et al., 2015; Spartano et al., 2019). Similar findings have been observed in studies investigating an association between daily activity or sedentary bout length and incident dementia (Buchman et al., 2012; Del Pozo Cruz, Ahmadi, Naismith, & Stamatakis, 2022; Raichlen et al., 2023) or in studies of fewer than 50 individuals with prevalent dementia (Hartman, Karssemeijer, van Diepen, Olde Rikkert, & Thijssen, 2018; Hatfield, Herbert, van Someren, Hodges, & Hastings, 2004). These findings suggest that accelerometry devices could serve as biometric markers of activity-related symptoms and cognitive decline and/or dementia in older adults, but no large, nationally representative studies have tackled the relationship between prevalent dementia and activity.

Likewise, only a few studies have attempted to use accelerometers to differentiate between different dementia subtypes. Given that differentiating dementia with Lewy bodies (DLB) from Alzheimer’s disease (AD) is challenging (Morra & Donovick, 2014; Vann Jones & O’Brien, 2014) and that motor disturbances are more common in DLB than in AD, accelerometers that objectively measure the motor domain between clinic visits could help improve the identification of early dementia.

One large-scale effort to make accelerometry data available to researchers is the National Health and Aging Trends Study (NHATS), which collects a range of accelerometer-derived measures and demographic, behavioral, and health-related covariates in older adults (Freedman, Schrack, Skehan, & Kasper, 2022). We seek to use the NHATS accelerometry data to better understand differences in PA and activity fragmentation between individuals with and without dementia and to examine how these differences change with age. In addition, we will make similar comparisons in a smaller sample of older adults with DLB and AD from the Technology for Early Dementia Diagnosis (TEDD) study to assess the utility of using accelerometer-derived metrics to identify PA-related differences across the two groups.

## Methods

### NHATS study population

Data were obtained from NHATS, a large consortium that conducts in-person interviews on the physical, cognitive, and sensory capacity of a nationally representative sample of U.S. Medicare beneficiaries who are age 65 or older (Freedman & Kasper, 2019). NHATS participants are classified as having probable, possible, or no dementia per a report by the participant or proxy respondent that a doctor told the participant that he/she had dementia or Alzheimer’s disease, a score that indicates probable dementia on the AD8 Dementia Screening Interview (Galvin et al., 2005; Galvin, Roe, Xiong, & Morris, 2006), or cognitive tests that evaluate the sample person’s memory, orientation, and executive function (Kasper, Freedman, & Spillman, 2013). In this analysis, participants with probable or possible dementia were labeled as having dementia.

In Round 11 (2021), NHATS began collecting PA data from a subsample of the NHATS cohort (n = 872). After an interview with a study team member, participants were instructed to wear the ActiGraph CentrePoint Insight Watch (a triaxial accelerometer) on their nondominant wrist for 7 days after the interview day (i.e., 8 days in total). The participants were to wear the devices at all times, except when swimming or bathing for more than 30 minutes. The devices were set to a sampling rate of 64 Hz.

Raw accelerometer data files were unavailable for NHATS participants. Non-wear time was estimated as any interval of 90 minutes or longer, and a valid day was defined as wear time greater than 90% or 1296 minutes per day (Schrack, Skehan, & Freedman, 2023). Of the 872 participants who were eligible to wear the activity watch, 747 (86%) returned an activity watch with at least 1 day of valid data. We excluded participants who had ≤ 3 days of valid data (n = 9) given that these individuals exhibited activity outcomes that were highly variable. This resulted in an analytic sample size of 738 NHATS participants, of which 104 had dementia and 634 did not.

### Technology for Early Dementia Diagnosis (TEDD) study population

Participants with DLB (n=12) and AD (n=8) were recruited from VA Puget Sound Health Care System (VA Puget Sound) and the Puget Sound community as part of a larger NIH-funded study to explore how novel measurements of activity, sleep, cognition, and behavior could be used to differentiate individuals with DLB from individuals with AD. All participants provided informed consent and were enrolled in a protocol approved by the VA Puget Sound institutional review board (IRB). All participants either met criteria for probable DLB according to the Fourth Consensus Report (McKeith et al., 2017) or for probable AD dementia according to the National Institute on Aging–Alzheimer’s Association Workgroup criteria (McKhann et al., 2011).

After receiving a brief tutorial from a study coordinator (either remotely or in person), participants were instructed to wear ActiGraph’s triaxial wGT3X-BT accelerometry device on their nondominant wrist for 14 consecutive full days and nights; this included taping the watch in place so that it could not be removed during the study. The participants were to wear the devices at all times, including when bathing or swimming. The devices were set to a sampling rate of 30 Hz.

Raw accelerometer data files for each participant were processed in R (version 4.1.1). At the beginning of the study, some participants began wearing the devices shortly after the recording had started, and on the final day of recording, some participants removed the devices early. These partial non-wear days were identified using the data quality visualization feature of the GGIR package (version 2.8.2) and were subsequently removed from the analysis; no other non-wear times were identified in any participants. Across all participants, 10 days of partially recorded data were identified and removed. Each participant ultimately had at least 12 days of accelerometer data with complete wear time.

### Generation of activity outcomes

Accelerometer data from both data sources were presented as the number of vector magnitude activity counts occurring in each second of wear time. We generated three primary outcome measures for each data source: mean daily activity counts, mean active bout length, and mean resting bout length. To calculate average daily counts, we summed the number of counts occurring each day and then averaged the number of daily counts over all valid days of wear time for each person individually. The fragmentation outcomes (mean active bout length and mean resting bout length) were provided by NHATS and are defined as the average duration (in minutes) of both active bouts (≥ 1,853 activity counts per minute) and resting bouts (< 1,853 activity counts per minute). We created these outcomes manually in the TEDD sample by classifying each minute of wear time as active or resting using the same cutoff that NHATS uses (i.e., 1,853 activity counts per minute) and calculating the average duration of each activity class for each participant.

### Statistical analyses

For NHATS analyses, survey weighted linear regression models were run using the svyglm function of the survey package (version 4.2-1) in R. The models were weighted using the survey weights provided by NHATS. Three models were created: one with daily activity counts as the outcome, one with active bout length as the outcome, and one with resting bout length as the outcome. The predictor variables in these models included race (binary; white vs. non-white), sex (binary; male vs. female), age, dementia status (binary; dementia vs. no dementia), and an interaction term between age and dementia status.

For TEDD analyses, multiple linear regression models were run with the same outcomes as the NHATS models. Predictors in these models included age and dementia subtype (binary; DLB vs. AD). The race and sex terms were dropped from these models given that the sample had limited variability in race and sex and that the sample size was likely too small to support more complex modeling. We compared the association between age and all three outcomes across the two dementia-related subgroups of each sample. Additionally, we compared the predicted value of all three outcomes at the mean age of each sample (i.e., 79 for NHATS and 74 for TEDD).

As described above, NHATS and TEDD used different ActiGraph triaxial accelerometer models that were set to record at different sampling rates and for different amounts of time; they applied different standards regarding non-wear time; and they likely used different processing methods to convert raw accelerometry data into the outcome measures in this study. These differences prevented us from directly comparing measures of daily activity, active bout length, and resting bout length between the two cohorts.

## Results

In the NHATS sample, participants with dementia were slightly older, more likely to be female, and more likely to be nonwhite than participants without dementia (see Table 1). In the TEDD sample, participants with DLB were slightly younger than participants with AD, but the two groups were similar in sex and race (see Table 2). The TEDD participants were also much more likely to be male (90%) than the NHATS participants (48.5%); this finding was not surprising given that the TEDD study recruited primarily from a population of Veterans.

**Table 1:** Demographic summary of NHATS participants with and without dementia.

| Variable | Dementia<br>(n=104) | No dementia<br>(n=634) | Overall<br>(n=738) |
| --- | --- | --- | --- |
| Age |  |  |  |
| Mean (SD) | 81.9 (6.2) | 78.8 (5.77) | 79.2 (5.9) |
| Median (min, max) | 81 (72, 100) | 77 (71, 100) | 78 (71, 85) |
| Sex |  |  |  |
| Male | 43 (41.1%) | 295 (46.5%) | 338 (45.8%) |
| Race |  |  |  |
| White | 67 (64.4%) | 526 (83%) | 593 (80.4%) |

**Table 2:**
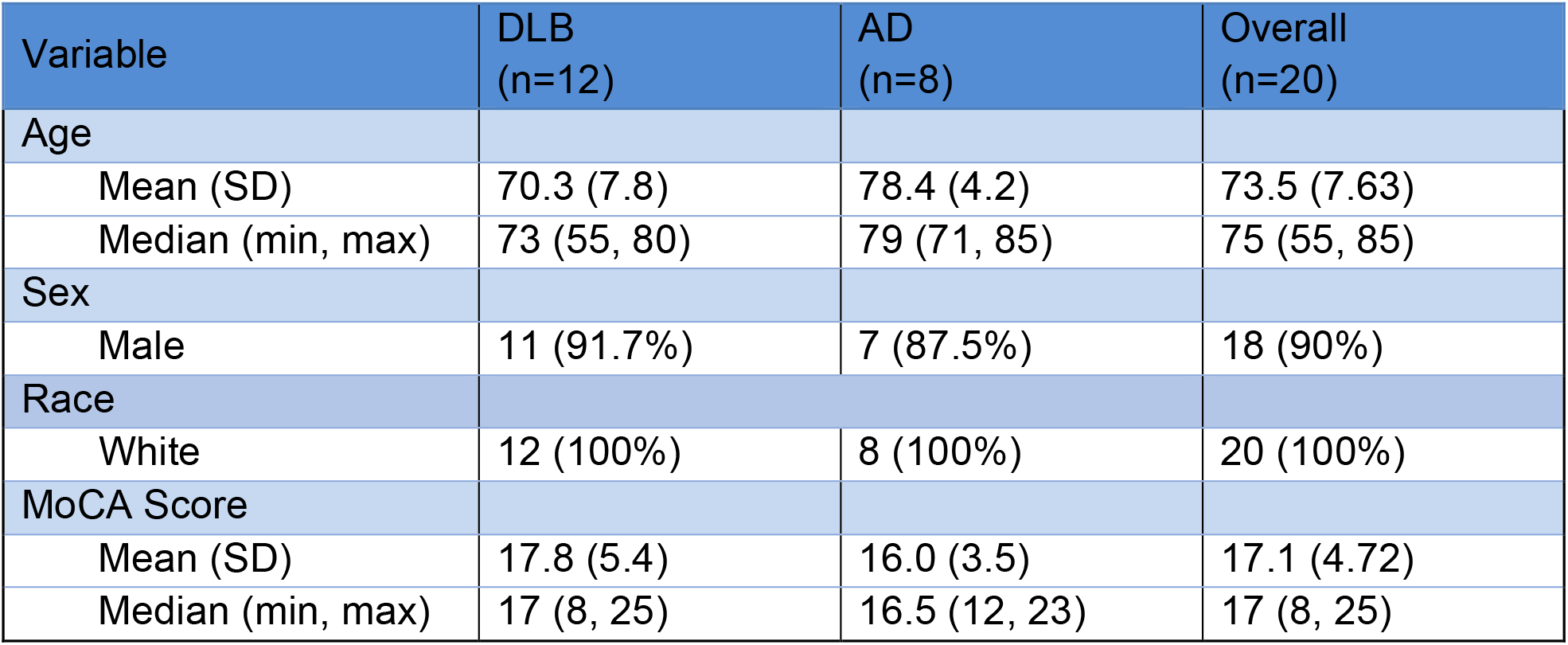
Demographic and cognitive summary of TEDD participants with AD and DLB.

| Variable | DLB<br>(n=12) | AD<br>(n=8) | Overall<br>(n=20) |
| --- | --- | --- | --- |
| Age |  |  |  |
| Mean (SD) | 70.3 (7.8) | 78.4 (4.2) | 73.5 (7.63) |
| Median (min, max) | 73 (55, 80) | 79 (71, 85) | 75 (55, 85) |
| Sex |  |  |  |
| Male | 11 (91.7%) | 7 (87.5%) | 18 (90%) |
| Race |  |  |  |
| White | 12 (100%) | 8 (100%) | 20 (100%) |
| MoCA Score |  |  |  |
| Mean (SD) | 17.8 (5.4) | 16.0 (3.5) | 17.1 (4.72) |
| Median (min, max) | 17 (8, 25) | 16.5 (12, 23) | 17 (8, 25) |

Although the NHATS collects cognitive data, specific scores on cognitive assessments were not available for this analysis. In the TEDD sample, participants with DLB performed slightly worse on the Montreal Cognitive Assessment (MoCA) than TEDD participants with AD (see Table 2) despite being slightly younger.

Mean daily activity counts in the NHATS sample ranged from 0.23 million to 4.29 million. For the NHATS participants with and without dementia, the mean (SD) daily activity counts were 1.33 (0.57) million and 1.73 (0.62) million, the mean (SD) sedentary bout durations were 6.60 (3.01) minutes and 5.73 (1.68) minutes, and the mean (SD) active bout durations were 7.34 (3.34) minutes and 8.47 (3.79) minutes, respectively (Figure 1).

**Figure 1:**
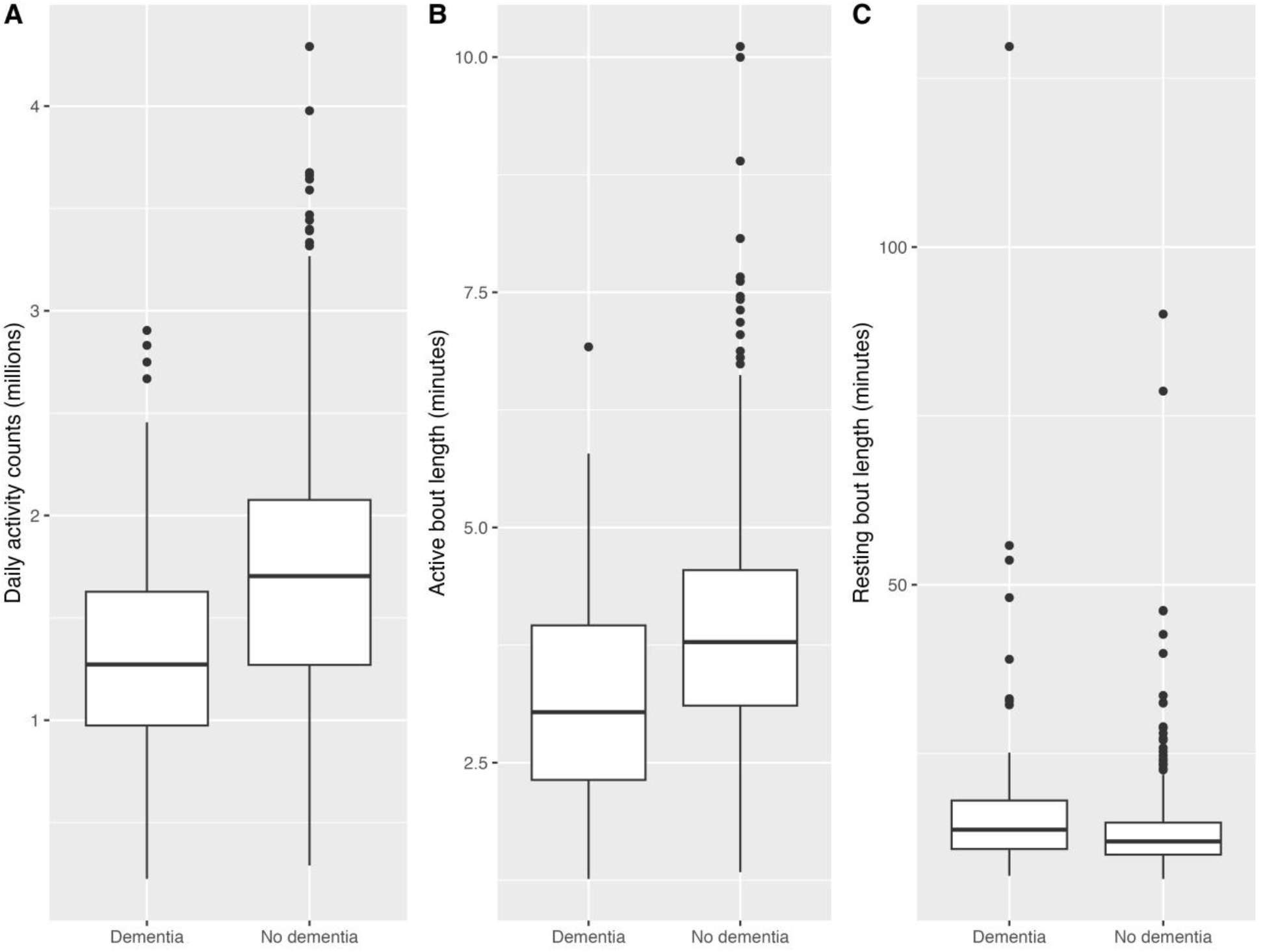
Boxplots of daily activity counts, active bout length, and resting bout length for NHATS participants with and without dementia.

Mean daily activity counts in the TEDD sample ranged from 0.38 million to 3.03 million. For the TEDD participants with DLB and with AD, the mean (SD) daily activity counts were 1.38 (0.69) million and 1.62 (0.23) million, the mean (SD) sedentary bout durations were 19.2 (12.8) minutes and 12.3 (2.91) minutes, and the mean (SD) active bout durations were 6.44 (2.56) minutes and 7.06 (1.51) minutes, respectively (Figure 2).

**Figure 2:**
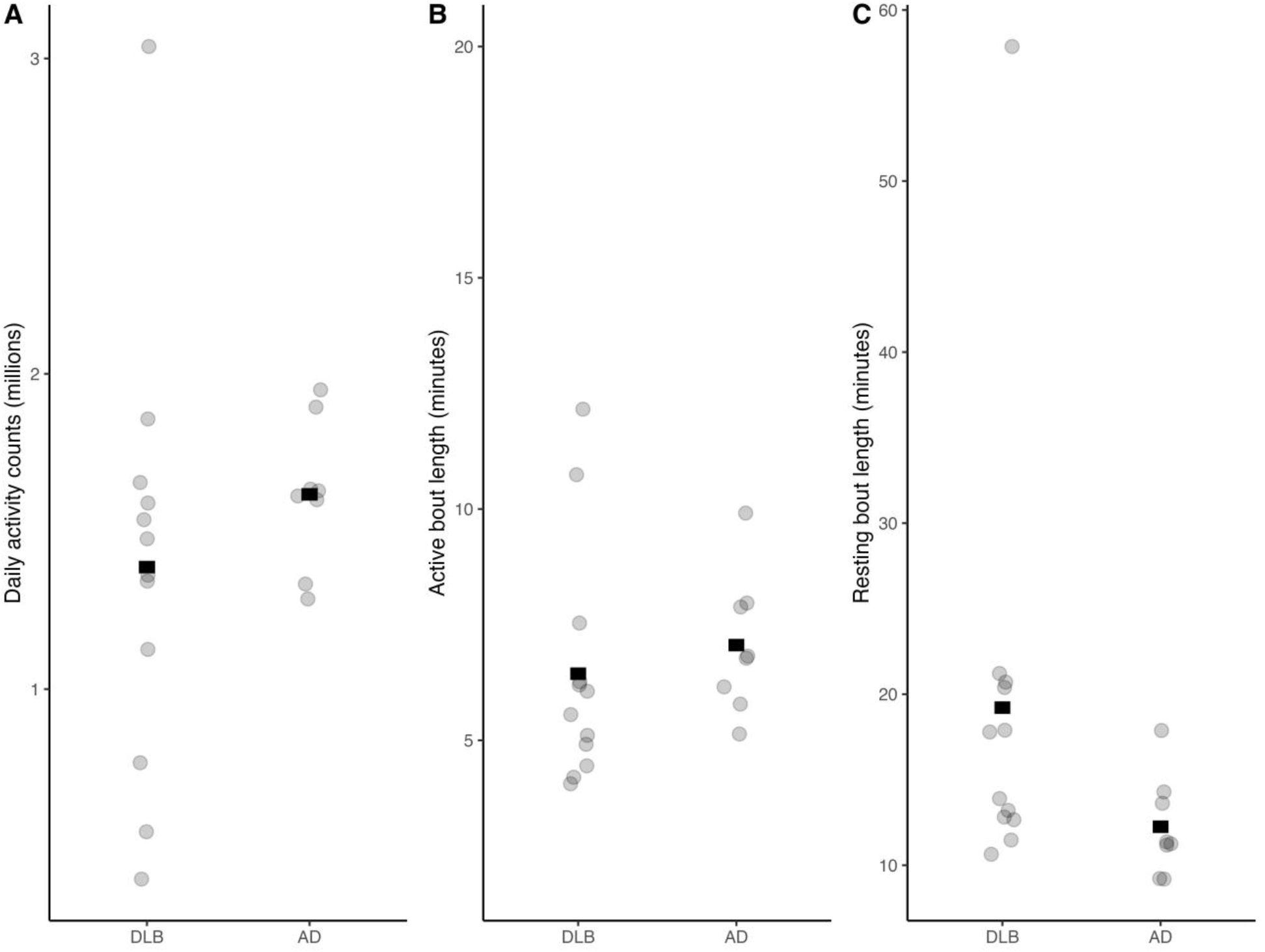
Scatterplots with bars indicating the mean daily activity counts, active bout length, and resting bout length for TEDD participants with DLB and AD.

In our NHATS models, a one-year difference in age tended to be associated with less daily activity for both participants with dementia (-0.015 [95 % CI -0.030, 0.001] million activity counts) and participants without dementia (-0.032 [95 % CI -0.039, -0.024] million activity counts), with shorter active bout lengths for both participants with dementia (-0.012 [95 % CI -0.044, 0.019] minutes) and participants without dementia (-0.048 [95 % CI -0.063, -0.032] minutes), and with a longer resting bout length for both participants with dementia (0.278 [95 % CI -0.033, 0.589] minutes) and participants without dementia (0.192 [95 % CI 0.066, 0.318] minutes) (Table 3). Participants without dementia exhibited significantly higher daily activity counts (0.406 million activity counts [95% CI 0.256 million, 0.555 million]), a longer active bout length (0.78 minutes [95% CI 0.48, 1.07]), and a shorter resting bout length (-3.25 minutes [95% CI –6.21, -0.29]) at the mean sample age of 79 (Table 5).

**Table 3:**
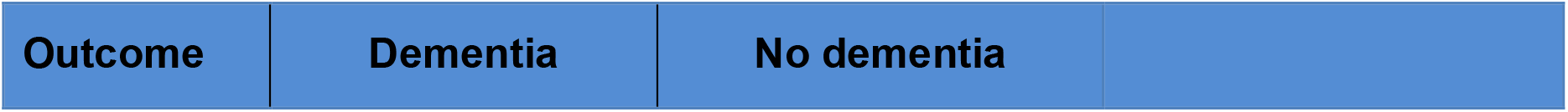
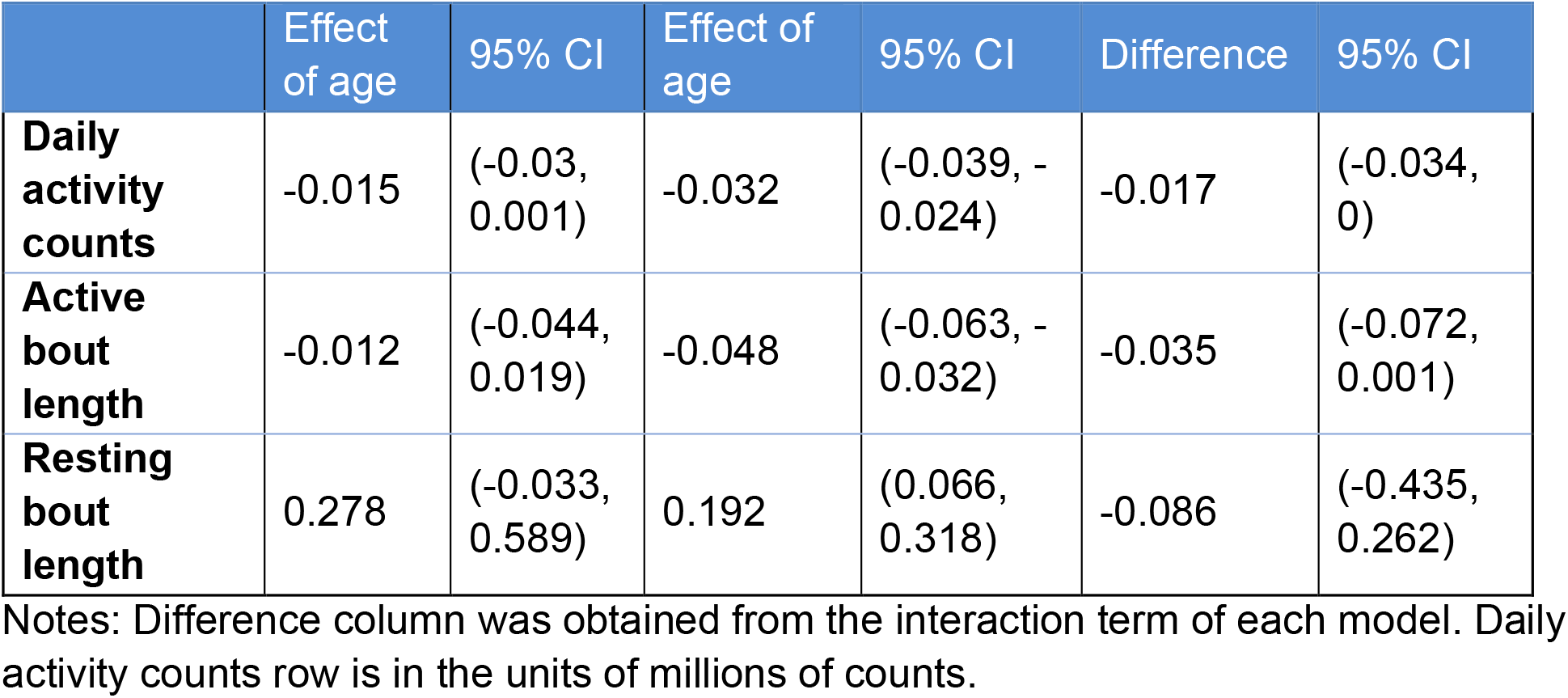
The effect of a one-year difference in age on daily activity counts, active bout length, and resting bout length for NHATS participants with and without dementia.

|  | Effect of age | 95% CI | Effect of age | 95% CI | Difference | 95% CI |
| --- | --- | --- | --- | --- | --- | --- |
| **Daily activity counts** | -0.015 | (-0.03, 0.001) | -0.032 | (-0.039, -0.024) | -0.017 | (-0.034, 0) |
| **Active bout length** | -0.012 | (-0.044, 0.019) | -0.048 | (-0.063, -0.032) | -0.035 | (-0.072, 0.001) |
| **Resting bout length** | 0.278 | (-0.033, 0.589) | 0.192 | (0.066, 0.318) | -0.086 | (-0.435, 0.262) |

Notes: Difference column was obtained from the interaction term of each model. Daily activity counts row is in the units of millions of counts.

In our TEDD models, a one-year difference in age was associated with slightly less daily activity (-0.005 [-0.05, 0.04] million activity counts) for participants with DLB and slightly more daily activity (i.e., 0.003 [-0.107, 0.1] million activity counts) for participants with AD, with a longer active bout length (0.017 [-0.161, 0.195] minutes) for participants with DLB and a shorter active bout length (-0.071 [-0.479, 0.337] minutes) for participants with AD, and with a longer resting bout length (0.396 [-0.399, 1.191] minutes) for participants with DLB (Table 4) and a shorter resting bout length (-0.101 [-1.925, 1.723] minutes) for participants with AD. There were no statistically significant differences between participants with DLB and participants with AD in daily activity counts (-0.262 million activity counts difference [95% CI - 1.032 million, .508 million]), active bout length (-0.866 minute difference [95% CI –3.926, 2.194]), or resting bout length (8.005 minute difference [95% CI –5.668, 21.678]) at the mean sample age of 74 (Table 6).

**Table 4:** The effect of a one-year difference in age on daily activity counts, active bout length, and resting bout length for TEDD participants with AD and DLB.

| Outcome | DLB |  | AD |  |  |  |
| --- | --- | --- | --- | --- | --- | --- |
|  | Effect of age | 95% CI | Effect of age | 95% CI | Difference | 95% CI |
| **Daily activity counts** | -0.005 | (-0.05, 0.04) | -0.003 | (-0.107, 0.100) | 0.002 | (-0.111, 0.114) |
| **Active bout length** | 0.017 | (-0.161, 0.195) | -0.071 | (-0.479, 0.337) | -0.088 | (-0.533, 0.358) |
| **Resting bout length** | 0.396 | (-0.399, 1.191) | -0.101 | (-1.925, 1.723) | -0.497 | (-2.486, 1.493) |

Notes: Difference column was obtained from the interaction term of each model. Daily activity counts row is in the units of millions of counts.

**Table 5:**
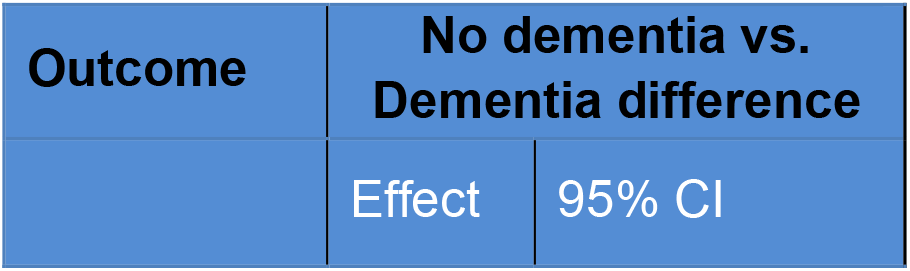
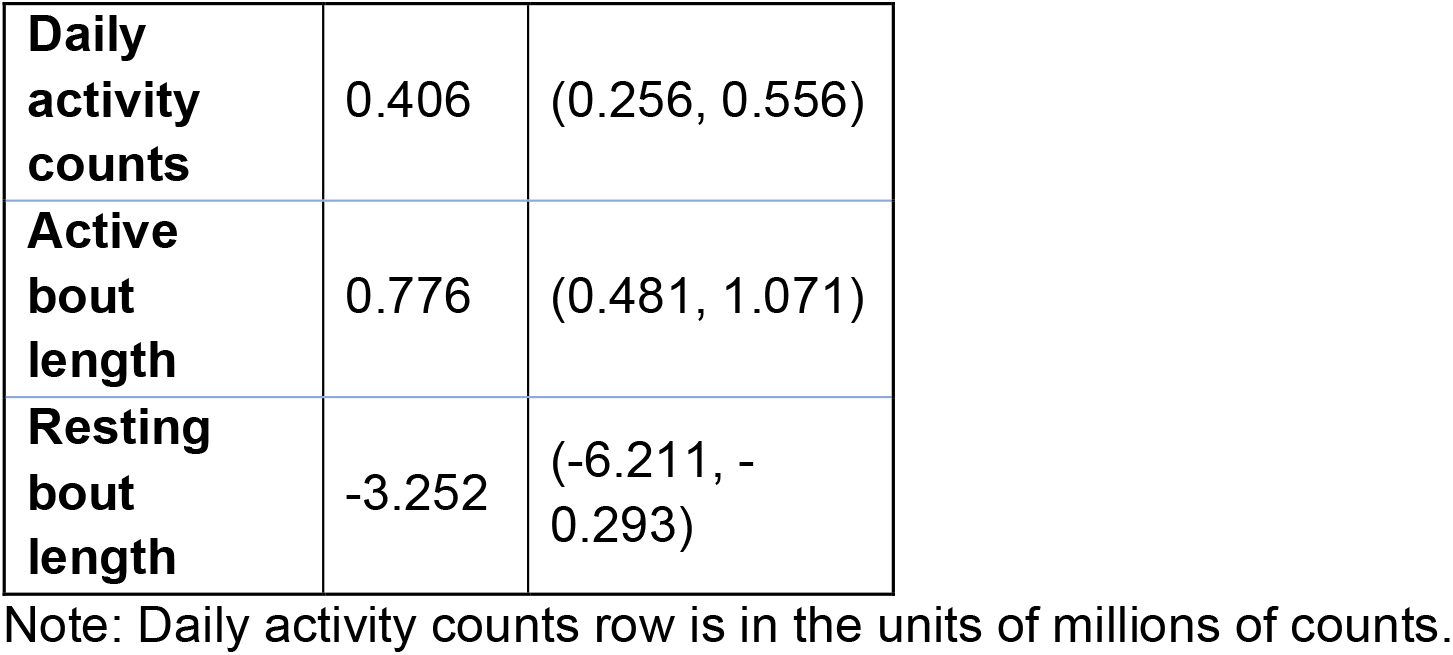
The difference in predicted daily activity counts, active bout length, and resting bout length for NHATS participants with and without dementia at the mean age of the sample (i.e., age 79)

| Outcome | No dementia vs. Dementia difference |  |
| --- | --- | --- |
|  | Effect | 95% CI |
| **Daily activity counts** | 0.406 | (0.256, 0.556) |
| **Active bout length** | 0.776 | (0.481, 1.071) |
| **Resting bout length** | -3.252 | (-6.211, -0.293) |

Note: Daily activity counts row is in the units of millions of counts.

**Table 6:** The difference in predicted daily activity counts, active bout length, and resting bout length for TEDD participants with AD and DLB at the mean age of the sample (i.e., 74)

| **Outcome** | **DLB vs. AD difference** |  |
| --- | --- | --- |
|  | **Effect** | **95% CI** |
| **Daily activity counts** | -0.262 | (-1.032, 0.508) |
| **Active bout length** | -0.866 | (-3.926, 2.194) |
| **Resting bout length** | 8.005 | (- 5.668, 21.678) |

Note: Daily activity counts row is in the units of millions of counts.

As noted in the Methods section, differences in data collection and processing prevented us from directly comparing the NHATS and TEDD participants on daily activity, active bout length, and resting bout length.

## Discussion

To our knowledge, this is the first study to use the NHATS dataset to compare activity and sedentary behavior in individuals with and without dementia. We found that individuals with dementia had less daily activity counts, a shorter active bout length, and a longer resting bout length than individuals without dementia. These findings, which were identified in a large, nationally representative sample, are consistent with the findings of other, smaller studies of those with and without dementia (Hartman et al., 2018; Mc Ardle et al., 2020; van Alphen et al., 2016), as well as studies of incident dementia (Buchman et al., 2012; Del Pozo Cruz et al., 2022; Raichlen et al., 2023). Given that both age and dementia status likely contribute to differences in activity, we were particularly interested in evaluating how age affected the relationship between our accelerometry measures and dementia status. We identified a negative association between age and both daily activity counts and active bout length such that, as one might expect, older NHATS participants were less active than younger NHATS participants. We also found that these negative associations were smaller for NHATS participants with dementia than NHATS participants without dementia (see Figure3a, Figure 3b, Table 3). This difference might be related to the fact that daily activity counts and active bout length values were low enough among NHATS participants with dementia that the older individuals with dementia were not markedly less active than the younger individuals with dementia. In other words, the data from NHATS participants without dementia may suggest a normal age-related decline in total activity and active bout length, whereas the data from NHATS participants with dementia may suggest that activity levels and active bout length might decline precipitously sometime prior to the diagnosis of dementia and then remain consistently lower at all ages following dementia onset. To our knowledge, no studies have used accelerometry data to delineate this particular activity pattern in individuals with dementia, and longitudinal studies are thus needed in healthy individuals and individuals with mild cognitive impairment to observe activity differences during the early disease processes.

**Figure 3:**
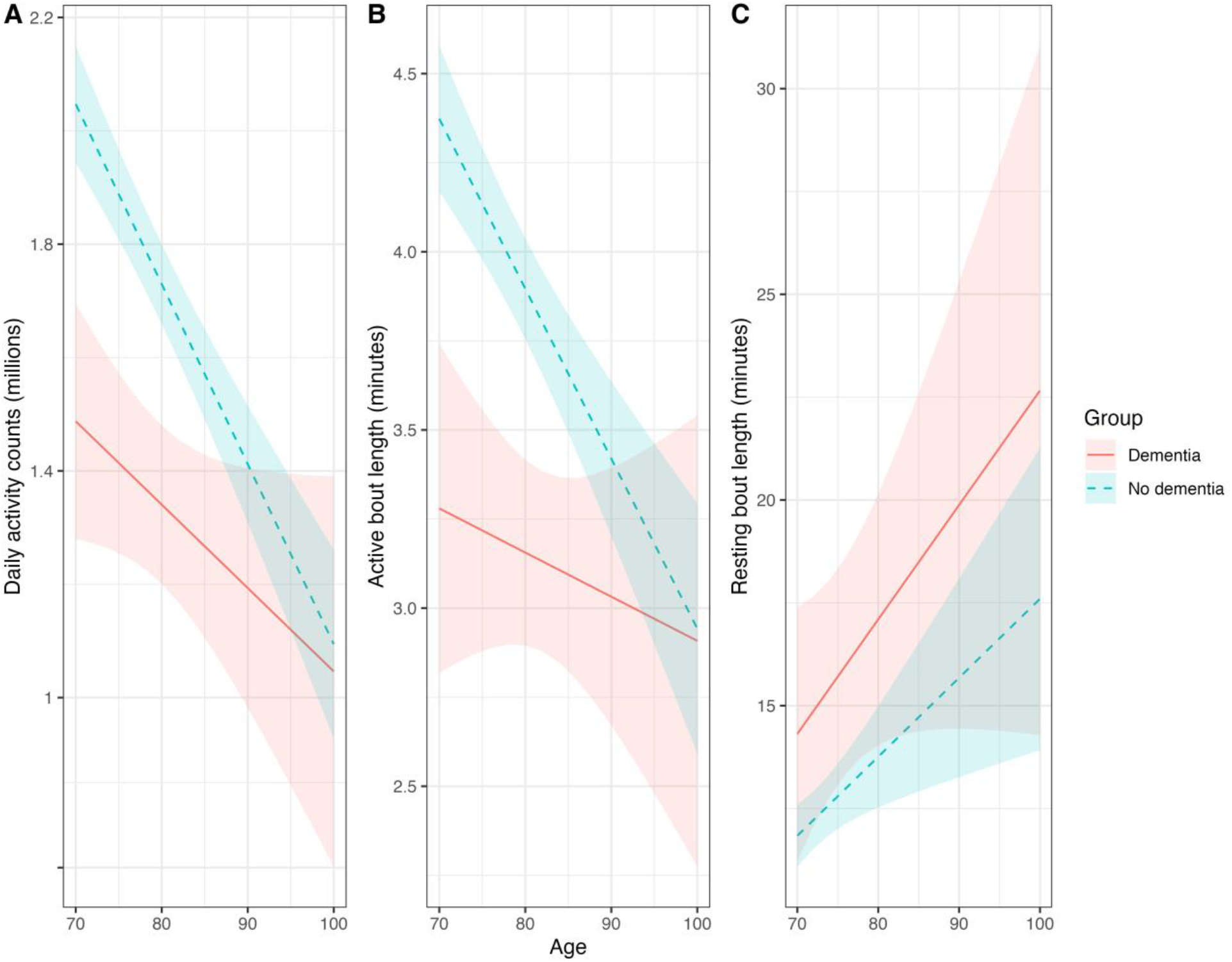
Predicted values of daily activity counts, active bout length, and resting bout length for NHATS participants with and without dementia by age.

**Figure 4:**
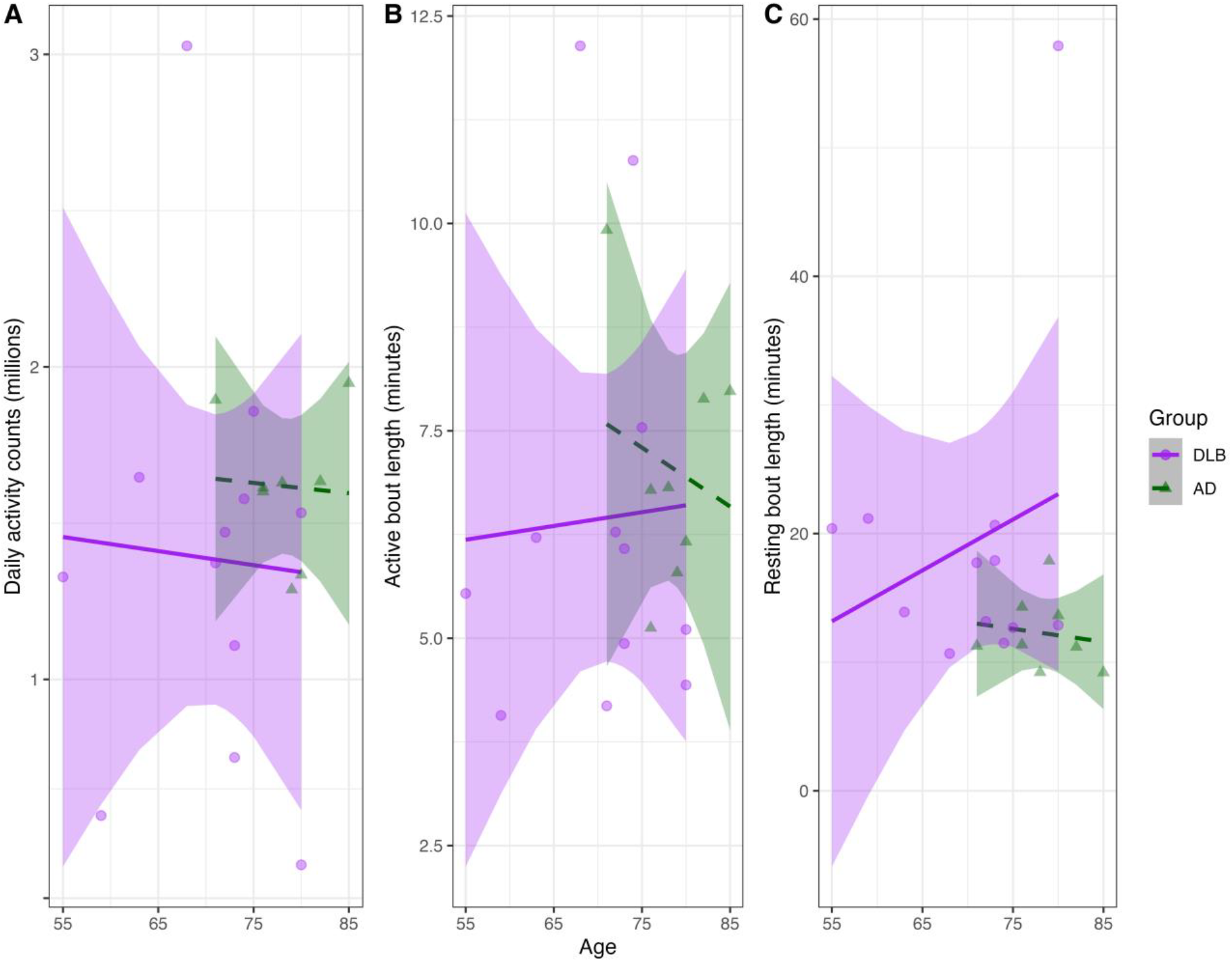
Predicted values of daily activity counts, active bout length, and resting bout length for TEDD participants with DLB and AD by age.

We did not observe a similar pattern for resting bout length, as NHATS participants with and without dementia exhibited similar age-related associations for resting bout length (see Figure 3c and Table 3). Our findings suggest that the effect of age on resting bout length may be more prominent than the effect of dementia on resting bout length. Although some accelerometry studies have identified sedentary activity as an outcome associated with prevalent dementia (Hartman et al., 2018) or incident dementia (Raichlen et al., 2023), other studies have had less convincing results. For example, in a study of self-reported leisure time (Raichlen et al., 2022), investigators found that spending more sedentary time watching television was associated with an increased risk of incident dementia, but spending more sedentary time on the computer was associated with a decreased risk of incident dementia, and in an accelerometry study, other investigators found no association between sedentary behaviors and incident dementia in a sample of older women (Nguyen et al., 2023). These inconsistent findings suggest that additional studies are needed to better understand how sedentary behaviors are related to dementia.

In addition to clarifying the role between dementia status, age, and accelerometry metrics, we also sought to identify specific differences between a small sample of TEDD participants with DLB and AD. Given that motor disturbances are more common in DLB than in AD (Yamada et al., 2020), we hypothesized that individuals with DLB would exhibit fewer total activity counts, shorter active bout lengths, and longer resting bout lengths than individuals with AD. The mean values for these three outcomes in our TEDD cohort were consistent with this hypothesis, but we found no statistically significant differences between the two groups. This suggests that studies with larger sample sizes in which age can be better matched between the groups may be able to establish more meaningful findings. We also observed no statistically significant differences in how the association between each outcome and age differed between the two groups (Table 4).

To our knowledge, McArdle et al. (2020) conducted the only other study to examine differences in accelerometer-derived outcomes across individuals with DLB (n = 30) and AD (n = 36). They found no statistically significant differences in the activity volume or patterns across groups while also observing a similar pattern such that the daily average values for minutes spent walking, step counts, and the number of active bouts were less in individuals with DLB than in individuals with AD. This suggests that studies with a larger sample size may be able to identify statistically significant differences between these two groups.

Perhaps the most striking difference in the accelerometry measures between our participants with DLB and AD, then, was that participants with DLB exhibited much greater variability in each outcome than participants with AD (Figure 2). Future accelerometry research that focuses specifically on individuals with DLB should investigate whether this variability is the result of randomness or something inherent to DLB.

An important limitation of this study is that differences in accelerometers, sampling rates, and processing prevented us from directly comparing the performance of NHATS and TEDD participants with dementia. Likewise, because the NHATS does not require participants to complete a robust diagnostic process that might identify dementia subtypes, it is also not possible to characterize differences between DLB and AD using this sample. This meant that to understand potential differences between dementia subtypes, we had to rely on the much smaller TEDD sample. There, a limitation is the sample size and age range. Given that the DLB group was on average younger than the AD group and that age has an established association with activity-based outcomes, future studies should concentrate on having participants with DLB and AD who are better matched on age. Lastly, both the NHATS and TEDD accelerometer data were collected during the COVID-19 pandemic, and we do not have a full understanding of how that context may have affected activity levels and sedentary behavior.

In conclusion, we used a large survey-based sample of individuals with and without dementia to identify differences in daily activity, active bout length, and resting bout length. These findings suggest that accelerometers may have the potential to help detect dementia, which may be particularly useful between clinical visits. The use of daily activity and active bout length measures may be especially useful in this regard, given that the association between these outcomes and dementia status was prominent, notably at the sample’s younger ages. We also extended these analyses to a smaller sample of dementia subtypes, and although we did not have a sufficient sample size to identify statistically significant differences between individuals with DLB and AD, our findings suggest that there is potential for accelerometers to aid in differentiating these two neurodegenerative diseases.

## Data Availability

Some data used in the present study are available online at https://nhats.org and other data are available upon reasonable request to the authors.

